# Almitrine as a non ventilatory strategy to improve intrapulmonary shunt in COVID-19 patients

**DOI:** 10.1101/2020.05.18.20105502

**Authors:** MR Losser, C Lapoix, B Champigneulle, M Delannoy, JF Payen, D Payen

**Author notes:** e-mails: Marie Reine Losser; Coline Lapoix; Benoit Champigneulle; Matthieu Delannoy; Jean-Francois Payen; Didier Payen.

## Abstract

In severe COVID-19 pulmonary failure, hypoxia is mainly related to pulmonary vasodilation with altered hypoxic pulmonary vasoconstriction (HPV). Besides prone positioning, other non-ventilatory strategies may reduce the intrapulmonary shunt. This study has investigated almitrine, a pharmacological option to improve oxygenation.

Patients and Method. A case control series of 17 confirmed COVID-19 mechanically ventilated patients in prone or supine positioning was collected: 10 patients received two doses of almitrine (4 and 12 mcg/kg/min) at 30-45 min interval each, and were compared to 7 “control” COVID-matched patients conventionally treated. The end-point was the reduction of intra-pulmonary shunt increasing the PaO_2_ and ScvO_2_.

Results Patients were male (59%) with median (25^th^, 75th percentiles) age of 70 (54-78) years and a BMI of 29 (23-34). At stable mechanical ventilatory settings, PaO_2_ (mmHg) at FiO_2_ 1 (135 (85, 195) to 214 (121, 275); p = 0.06) tended to increase with almitrine. This difference was significant when the best PaO_2_ between the 2 doses was used: 215 (123,294) vs baseline (p = 0.01). A concomitant increase in ScvO_2_ occurred ((73 (72, 76) to 82 (80, 87); p = 0.02). Eight over 10 almitrine-treated patients increased their PaO_2_, with no clear dose-effect. During the same time, the controls did not change PaO_2_.

In conclusion, in early COVID-19 with severe hypoxemia, almitrine infusion is associated with improved oxygenation in prone or supine positioning. This pharmacological intervention may offer an alternative and/or an additional effect to proning and might delay or avoid more demanding modalities such as ECMO.

## Introduction

The clinical presentation of COVID-19 disease is heterogenous, ranging from no symptoms to severe acute respiratory failure (ARF), which may have a poor prognosis. (1, 2) The lung contamination by SARS-CoV-2 is typically characterized by a major difficulty to oxygenate the arterial blood. (3) This severe hypoxemia is associated with preserved respiratory mechanical properties, in particular the pulmonary system compliance. As recently quoted by Gattinoni et al(4), the COVID-19 pneumonia seems to have 2 different phenotypes: the early phase, with severe hypoxemia and close to normal respiratory mechanics with a moderate effect of PEEP on lung recruitment; the later phase, corresponding to the more “classic” ARDS alteration of respiratory mechanics such as a reduced compliance with chest CT Scan images of diffuse ground-glass opacities and condensations. The hypoxia during the early phase seems to mainly result from an important ventilation/perfusion (VA/Q) mismatch(5) associated with an altered pulmonary vasoconstriction. The “protective” mechanism called hypoxic pulmonary vasoconstriction (HPV) normally reduces the blood flow in poorly or non-ventilated areas towards aerated zones leading to reduce the (VA/Q) mismatch. HPV seems poorly functional in COVID-19 severe patients in absence of “cor pulmonale”.(2)

Most of the publications on severe COVID-19 describe intubated and mechanically ventilated patients, installed in prone positioning as an adjuvant therapy to limit hypoxia. The prone positioning in ARDS is supposed to facilitate alveolar recruitment, and to decrease the heterogeneity of compliance. (6) In theory, this modality should have a limited effect during the COVID-19 early phase. The recent publications demonstrated that the majority of patients improved PaO_2_ after prone positioning, (7) which suggests another mechanism to improve PaO_2_. The most probable was a pulmonary blood flow redistribution towards the ventilated areas by gravitational forces. This modality creates a huge work load to the care team, and increases the risk of endotracheal tube obstruction and malposition, and pressure sores on face and chest.(6)

In the 90s’, other non-ventilatory strategies have been reported including inhaled nitric oxide(8), almitrine bismesylate (9, 10) or a combination of both.(11) According to the French National agency for Drug Security (ANSM), only iv almitrine was indicated for hypoxic acute respiratory failure as Drug of Major Therapeutic Interest. The brutal COVID-19 outbreak with many severe hypoxic patients prompted intensivists to test iv almitrine to improve HPV(12, 13) and to reduce the intrapulmonary shunt. The use of ECMO is limited to trained centres, which could not meet COVID-19-related demands. We hypothesized that almitrine might restore even partially a HPV response, both in supine or prone positioning.

We investigated 10 COVID-19 patients mechanically ventilated at FiO_2_ 1 with a severe intrapulmonary shunt during their early phase. The emergency conditions and the acute high inflow of patients to ICU impeded the design of a randomized control trial. To eliminate the eventuality of a spontaneous evolution of hypoxia, these patients were compared with 7 control-matched COVID-19 patients treated conventionally.

### Patients and measurements

The Research Program database of the Departement d’Anesthésie-Reanimation Brabois Adulte was submitted to the “Direction de la Recherche et Innovation (ref 2020PI080)”, and was agreed by the research Ethical Committee (Saisine 263) of Centre Hospitalier Régional Universitaire (CHRU) de Nancy, France. The relatives or patients were questioned about objections to use almitrine and collected data for scientific purposes and/or potential publications or not. The statement including also the opposition forms was dated and recorded in medical file. The study was retrospectively registered at ClinicalTrials.govafter enrolment (NCT04380727, Principal Investigator: Marie-Reine LOSSER, M.D., Ph.D., May 7, 2020,

Between March16 and April 12, 2020, COVID-19 patients referred to ICU were screened to receive iv almitrine using the following criteria: a positive RT-PCR, a highly suggestive thoracic CTScan, and a severe hypoxemia leading to intubation for less than 3 days. The exclusion criteria were: the presence of an acute cor pulmonale on the trans-thoracic 2D Echo-Doppler(14) and abnormal liver function tests or hyperlactatemia.(15) Ten successive patients were studied according to the following protocol: baseline measurement in prone or supine positioning; second measurement 30 to 45 min after 4 mcg/kg/min iv almitrine bismesylate (Vectarion®, Servier Laboratory, Neuilly, France), and a third measurement 30-45 min after 12 mcg/kg/min infusion rate.(16) Because of a shortage of drug store at national level, a protocol using continuous infusion was not considered.

The recorded parameters were: the ventilatory settings unchanged along the protocol, including FiO_2_, PEEP level, tidal volume (VT), peak inspiratory pressure, plateau pressure; haemoglobin concentration, blood gases simultaneously sampled on the arterial catheter and on the central catheter to assess central venous oxygen saturation ScvO_2_, and arterial lactate, right atrial pressure; cardiac output when possible (Mostcare®, Vygon, Ecouen, France). Similar data were collected on a matched control group of COVID-19 patients, matched on gender, age, BMI and duration of mechanical ventilation, with serial measurements corresponding to the duration of almitrine testing. Data were reported according to the Strobe Statement for case control studies.(17)

### Statistical analysis

Data were reported as median (25th-75th percentiles) for continuous variables and as count (percentage) for categorical variables. Change in blood gases parameters during almitrine infusion were first assessed using a Friedman test followed by a pairwise Wilcoxon signed-ranks post-hoc test with Bonferroni correction when appropriate. Second, to perform a two-points comparison (i.e. baseline and after almitrine infusion) with the control group (baseline and H8 measurement), we only considered the best variation in term of PaO_2_ regardless of the infusion dose of almitrine (4 or 12 mcg/kg/min). In the two groups, difference between before (baseline) and after (almitrine infusion or H8) measurement was assessed using a Wilcoxon signed-ranks test. All tests were 2-sided and a p value <0.05 was considered as statistically significant. Statistical analyses were performed using R version 3.6.0 for Mac OS (The R Foundation for Statistical Computing, Vienna, Austria).

## Results

All patients had a confirmed diagnosis of COVID-19 (positive PCR testing). The median age was 70 (54-78) years, with 10/17 males, having a median BMI of 29 (23-34). Haemoglobin was stable at 12.2 (8.9-13.8) g/dL. Most of the patients were intubated just before or soon after ICU admission. Patients from both groups had co-morbidities, mostly metabolic and/or cardiovascular diseases under chronic treatment (Table 1). At the time of manuscript submission, in almitrine group 2 patients died and 6 patients were extubated and discharged from the ICU, the remaining being still mechanically ventilated. In the control group, 2 died and 5 were extubated and discharged from the ICU.

**Table 1:** clinical characteristics, comorbidities, chronic treatment, delay for almitrine testing and outcome. Top: almitrine COVID group; Bottom: “control” COVID group SAS = sleep apnea syndrome; NID = non-insulin-dependent diabetes; ECMO = extra-corporeal membrane oxygenation; COPD = chronic obstructive pulmonary disease; A Fib = atrial fibrillation. ED emergency department.

| *Patient* | *BMI* | *Past medical history* | *Medications* | *Delay for almitrine administration after intubation* | *Outcome* |
| --- | --- | --- | --- | --- | --- |
| 1 | 27 | A Fib – SAS | Proton pump inhibitor<br>Anti histaminic | D3 | Dead D17<br>Limitation of care |
| 2 | 26 | Ischemic heart disease – NIDD – prostate cancer | Aspirine<br>Calcium channel blocker<br>Fibrate/statins<br>ACE inhibitors<br>Proton pump inhibitor<br>Beta blocker<br>Benzodiazepine | D0 | Extubation D5<br>ICU discharge D19 |
| 3 | 31 | Hypertension – NIDD | □ blocker<br>Diuretic<br>Gliclazide<br>Aspirine | D4 | Extubation D6<br>ICU Discharge D7 |
| 4 | 29 | DVT – pulmonary and urinary tuberculosis | Oral anticoagulant | D1 | Extubation D3<br>Discharged home D8 |
| 5 | 35 | A Fib – Hypertension – NIDD - Transient stroke | Oral anticoagulant<br>Beta blocker<br>Antiarrhythmic type Ic<br>Sartan. Statins | D1 | Extubation D11<br>ICU discharge D15 |
| 6 | 23 | Myelodysplasia | 0 | D4 | Tracheotomy D18,<br>Dead D33,<br>Limitation of care |
| 7 | 33 | NIDD | Metformine | D1 | Extubated D13<br>ICU discharge D22 |
| 8 | 31 | Alcohol use disorder | 0 | D1 | ECMO D3 to D13,<br>still intubated at D30 |
| 9 | 23 | Hypertension – COPD – psoriasis | Local dermocorticoid<br>Aspirine<br>Proton pump inhibitor<br>benzodiazepine | D1 | Still intubated D18 |
| 10 | 24 | Hypertension – NIDD – ischemic stroke- depression | Insulin – Proton pump inhibitor – aspirine – Sartan – statins – Serotonin receptor inhibitor - | D 3 | Extubated at D8<br>ICU discharge D10 |

Table 1: clinical characteristics, comorbidities, chronic treatment, delay for almitrine testing and outcome. Top: almitrine COVID group; Bottom : “control” COVID group
| *Patient* | *BMI* | *Past medical history* | *Medications* | *Delay for blood gas after intubation* | *Outcome* |
| --- | --- | --- | --- | --- | --- |
| C1 | 31 | Hypertension – COPD | Diuretic – Sartan | D 4 | Extubated D14, ICU discharge D18 |
| C2 | 29 | Testicular carcinoma | 0 | D1 post intubation | Dead D24<br>Limitation of care |
| C3 | 30 | Breast cancer – obesity - depression | Benzodiazepine | D1 post intubation | Extubated D23<br>Stepdown care |
| C4 | 25 | Hypertension - smoking | NA | D1 post intubation | extubated D5, ICU discharge D7 |
| C5 | 32 | NIDD | Proton pump inhibitor<br>Metformine | D1 post intubation | Extubated D18 |
| C6 | 28 | Hypertension – NIDD –Prostate cancer – asthma | Aspirine – bronchodilator – beta blocker – proton pump inhibitor | D1 post intubation | Death D12<br>Limitation of care |
| C7 | 30 | Hypertension – IDD – COPD | Insulin – sartan – aspirine – statin | D1 post intubation | Extubated at D11<br>ICU discharge D16 |

Table 2 shows the hemodynamic, gas exchange and mechanical ventilation parameters from baseline, 4 and 12 mcg/kg/min of almitrine infusion. PaO_2_ tended to increase (p = 0.06) with a significant increase in ScvO_2_ (p =0.03). All other parameters did not change during the protocol. The changes in PaO_2_ did not parallel the increasing doses of almitrine (data not shown). In the control COVID group, none of the collected parameters changed after 8hrs (Supplementary table S1). The whiskers box plot of Figure 1 shows the median PaO_2_/FiO_2_ for almitrine and control groups. The best response in PaO_2_ increase starting from baseline was found significant only in the almitrine-treated group (p = 0.01), while it remained stable in controls. Figure 2 shows the individual data for both PaO_2_ and ScvO_2_ in both groups, which significantly increased only in the almitrine group. Eight over 10 patients increased their PaO_2_ with almitrine, with no clear relation with the level of infusion rate. Of note, when the drug was available to continue, the increase in PaO_2_ associated with almitrine infusion persisted (3 patients), which permitted to reduce the number of positioning changes. During this short perfusion time no hemodynamic side effects were observed and lactate remained stable (<1.5 mmol/L) (data not shown).

**Table 2:** Hemodynamic, pulmonary gas exchange and mechanical ventilation parameters at baseline and after 4 and 12 μg/kg/min of almitrine. BE = base excess; ScvO_2_ = central venous O_2_ saturation; Pra= right atrial pressure; CI = cardiac index; CO = cardiac output; Vt = tidal volume; RR = respiratory rate; PEEP = positive end-expiratory pressure; PIPressure = peak inspiratory pressure; Pplat = plateau pressure.

| *(median*<br>*(IQR))* | *Positioning* | *Baseline* | *4 µg/kg/min* | *12 µg/kg/min* | *p* |
| --- | --- | --- | --- | --- | --- |
| *n* |  | 10 | 10 | 10 |  |
| *FiO<sub>2</sub>* |  | 1.00 (1.00, 1.00) | 1.00 (1.00, 1.00) | 1.00 (1.00, 1.00) | **NA** |
| *condition (%)* | Prone | 3 (30.0) | 3 (30.0) | 3 (30.0) | **NA** |
|  | Supine | 7 (70.0) | 7 (70.0) | 7 (70.0) |  |
| *PaO<sub>2</sub> (mmHg)* |  | 135 (85, 195) | 149 (91, 28) | 215 (121, 275) | **0.06** |
| *PaCO<sub>2</sub> (mmHg)* |  | 42 (40, 45) | 42.1 (40, 45) | 43 (37, 46) | **0.06** |
| *pH* |  | 7.38 (7.36, 7.44) | 7.40 (7.36, 7.42) | 7.41 (7.36, 7.42) | **0.45** |
| *HCO<sub>3</sub> (mmol/L)* |  | 25.1 (23.5, 28.2) | 25.3 (24, 27.9) | 24.8 (23.9, 27.6) | **0.71** |
| *BE* |  | 1.3 (-1.4, 2.7) | 1.1 (-0.3, 2.4) | -0.3 (-1.5, 2.5) | **0.91** |
| *ScvO<sub>2</sub> (%)* |  | 73 (72, 76) | 81 (79, 82) | 85 (78, 87) * | **0.91** |
| *Pra (mmHg)* |  | 8 (7, 9) | 9 (8, 11) | 9 (8, 11) | **0.03** |
| *CI (l/min/m<sup>2</sup>)* |  | 2.1 (1.9, 2.2) | 2.2 (1.9, 2.3) | 2.2 (1.9, 2.5) | **0.33** |
| *CO (l/min)* |  | 4.2 (3.2, 4.3) | 4.1 (3.7, 4.6) | 4.3 (4.0, 4.5) | **0.42** |
| *Vt (ml)* |  | 425 (398, 465) | 425 (398, 465) | 425 (398, 465) | **0.42** |
| *RR (/min)* |  | 23 (22, 24) | 24 (22, 25) | 24 (22, 25) | **1** |
| *PEEP (cm H<sub>2</sub>O)* |  | 10 (10, 11) | 10 (8, 10) | 10 (10, 10) | **1** |
| *PIPressure (cm H<sub>2</sub>O)* |  | 27 (26, 29) | 27 (26, 30) | 27 (26, 29) | **0.14** |
| *Pplat (cm H<sub>2</sub>O)* |  | 22 (19, 24) | 22 (20, 24) | 22 (20, 24) | **1** |

**Figure 1:**
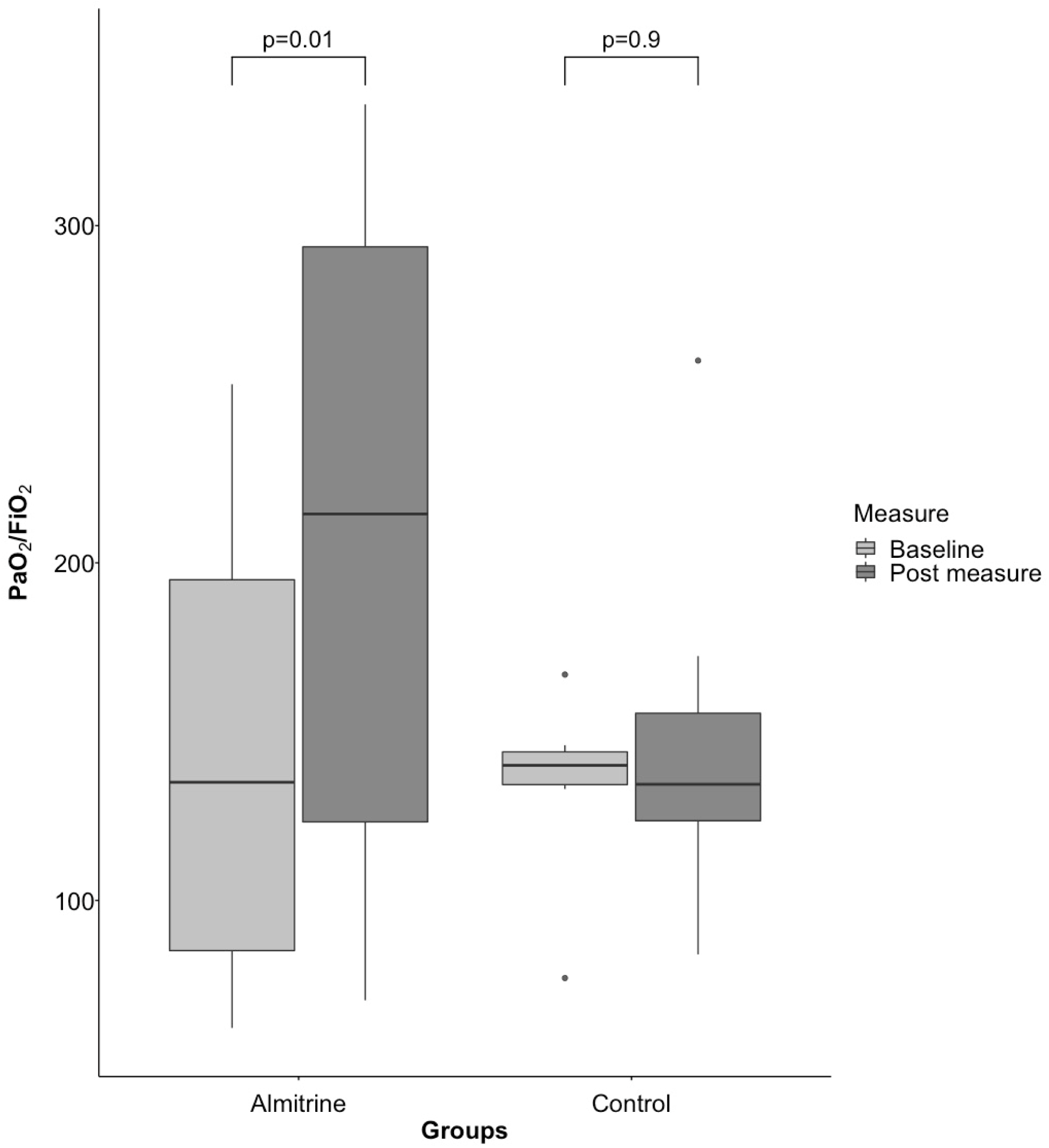
Boxplot representation of the PaO_2_/FiO_2_ ratio change in the almitrine group and in the control group between baseline and after treatment measurements (best dose response for patients treated with almitrine and 8-hours after baseline for matched control patients). *Tukey boxplots show median with 25^th^ and 75^th^ percentiles (lower and upper hinges). Whiskers extend from the correspondent hinge to the largest or smaller value not further than 1.5*interquartile range. Isolated points represent the outlier values*.

**Figure 2:**
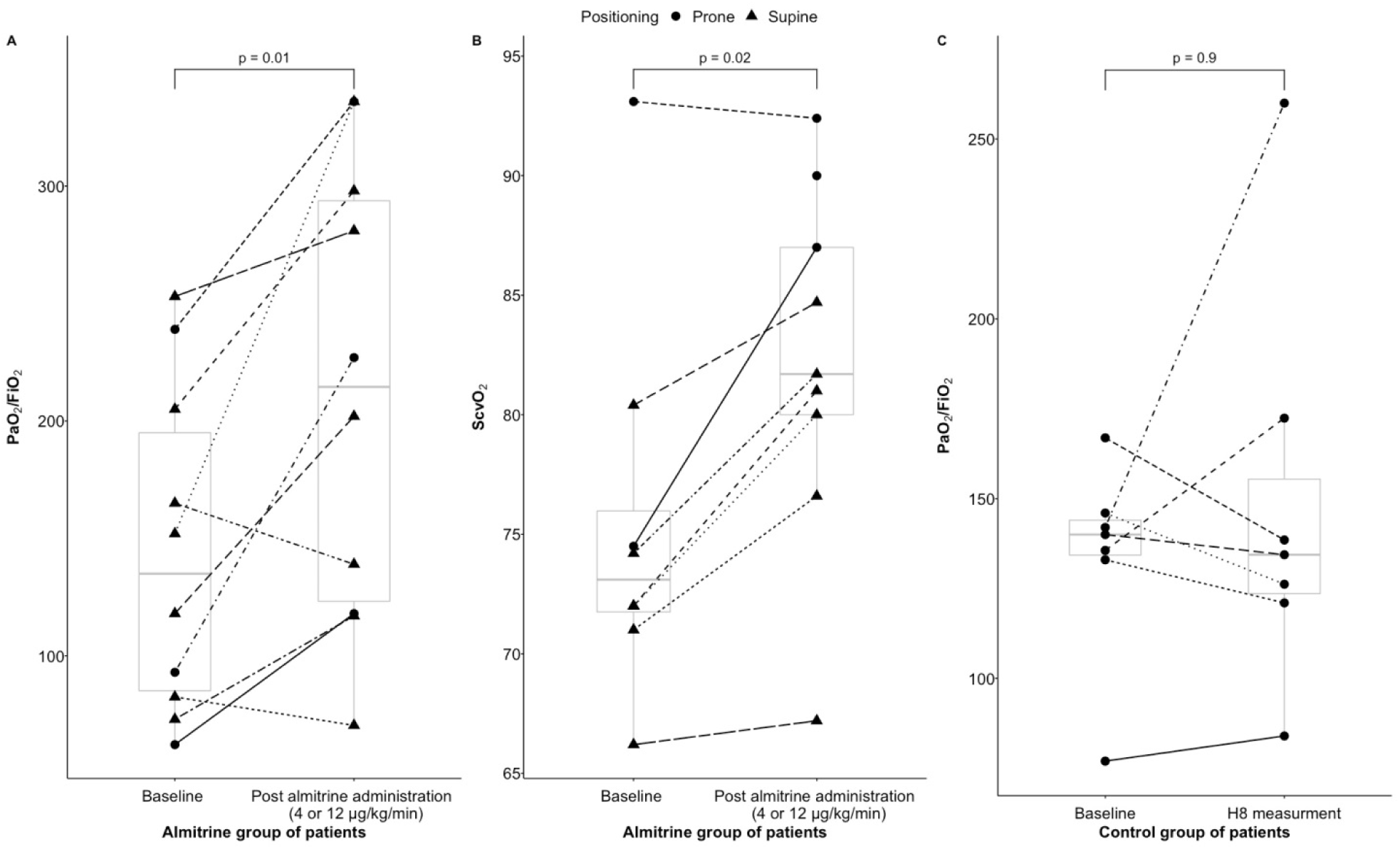
Individual change of the PaO_2_/FiO_2_ ratio (**A**) and the ScvO_2_ (**B**) between baseline and post almitrine administration (best dose response in term of PaO_2_/FiO_2_ ratio) in the almitrine group of patients (n=10). Individual change of the PaO_2_/FiO_2_ ratio (**C**) in the matched-control group (n=7) between baseline and H8. *ScvO_2_ measurements were not available for the control group*. *Shape of the individual point corresponds to the position of the patients (circle: prone positioning; triangle: supine positioning)*. *Tukey boxplots on the background show correspondent median with 25^th^ and 75^th^ percentiles (lower and upper hinges). Whiskers extend from the correspondent hinge to the largest or smaller value not further than 1.5*interquartile range*.

## Discussion

Intravenous almitrine was associated with almost a doubling of the PaO_2_/FiO_2_ ratio in the early phase of severe COVID-19 ARDS with no clear dose-effect relation. SvcO_2_ increased consistently, while neither Pra nor CI was altered. The individual responses related to almitrine infusion varied in amplitude between patients (fig2A), but 8 over 10 patients responded in term of PaO_2_ with an average of 80mmHg increase from baseline to the most efficient dose of almitrine.

In our experience, the patients admitted in ICU with a PaO_2_/FiO_2_ lower than 100 mmHg required a rescue intubation and mechanical ventilation at FIO_2_ close to 1. Standard ventilatory settings were used as previously reported in COVID-19(3), including moderate PEEP with a VT close to 6 ml/kg IBW. Among the non-ventilatory method to improve oxygenation, the prone positioning is leading with frequent and rapid increase in PaO_2_/FiO_2_.(6) As recently stated by ATS(6), prone positioning improves oxygenation mainly through respiratory mechanics improvement, limiting the heterogeneity of the gas volume partition. The apparent benefit of prone positioning on PaO_2_ in “early” COVID-19 with almost normal respiratory mechanics(18) strongly suggests that prone positioning improves hypoxia and reduces the intra-pulmonary VA/Q mismatch by another mechanism.

The absence of cor pulmonale despite severe ARF in COVID-19 indicates a relative dilatation of the pulmonary vascular bed with a “normal” pulmonary blood flow.(19) The logical hypothesis was that the prone positioning is changing the partition of pulmonary blood flow towards better ventilated areas by gravitation. The severe VA/Q mismatch in COVID-19 associated with pulmonary vasodilation indicates an alteration in HPV.(5) Years ago, we and others have shown that almitrine may spectacularly improve PaO_2_ by reducing the intra-pulmonary oxygen shunt.(5, 13) In the present pilot study, almitrine was used as a test to restore, even though partially, the HPV in supine or prone positioning (Fig 2B). The combination of these conditions (prone and almitrine) could then be seen as a combination of gravitational and pharmacological effects to improve the oxygen VA/Q mismatch. The observed increase in P/F ratio in 80% of the patients confirmed this approach. The small cohort of patients does not allow concluding about any additional effect of these therapies. Worthy of note, in 3 patients the re-positioning from prone to supine under almitrine prevented the loss of the benefit of prone positioning on PaO_2_ values. The observed results with almitrine would not be related to the spontaneous evolution of the patients as suggested by the stable PaO_2_ in the control COVID-19.

The PaO_2_ increase associated with almitrine infusion was concomitant with a significant increase in ScvO_2_. This observation confirms the absence of tissue hypoperfusion with no large peripheral O_2_ extraction, as suggested by the low lactate levels. This ScvO_2_ increase provides a greater reserve for O_2_ extraction in case of acute desaturation, and increased the level of dissolved O_2_, the diffusible form of oxygen to the tissues.(20)

This case control series has several limitations in addition to the small size cohort (17 patients). First, although the “control” COVID-19 patients were selected on stringent criteria to match the studied group, it was not randomized and could be biased. The stability in PaO_2_ during 8 hours reinforced the credence in an almitrine effect. In absence of a drug shortage, all of these patients would have received almitrine. This shortage resulted from the small national stock of almitrine facing an important demand during the pandemic. Second, although the data were prospectively collected, the overwhelmed care team capabilities in a dramatic context explains why some data were missing. Third, for the same reason, we were not able to administer the drug for a longer period than 36 to 48 hours for few patients. This precludes any conclusion about the potential benefit on mechanical ventilation duration and on the number of prone positioning. Following the same line of thinking, the almitrine test on arterial oxygenation cannot be proposed as either a prognostic test or a predicator of the prone position response.

In conclusion, in a case series of early hypoxemic COVID-19 pneumonia with acute respiratory failure, iv almitrine was associated with an improvement in arterial blood oxygenation both in prone or supine positioning in most of the patients, suggesting a partial recovery of the pulmonary vessels’ contractility. This pharmacological intervention may offer an alternative and/or an additional strategy to the prone positioning in severe COVID-19 ARDS. It may help to support the lung function when ECMO possibilities are very limited.

## Data Availability

all data referred to in the manuscript are available

## Authors contributions

MRL: study design, data collection, data interpretation, writing, literature search,

CL: data collection, figures,

MD: data collection, data interpretation

BC: figures, data analysis, data interpretation

JFP: data interpretation, writing, literature search

DP: literature search, study design, data analysis, data interpretation, writing

## Acknowledgements

We thank Doctors Marie Dominique Fratacci and Valérie Girard (Servier Laboratory, Courbevoie, France) for their permanent support and facilitation. We thank Drs Béatrice Demoré and Nathalie Commun from Pharmacie CHRU Nancy for permanent support.

## Major findings

During COVID-19, pulmonary blood vessel dilatation induces a severe intrapulmonary oxygen shunt. Almitrine *iv* infusion was associated with a rapid and significant improvement of PaO_2_ in mechanically ventilated patients during both prone or supine positioning.

